# Microvascular Thrombosis and Acute Kidney Injury in COVID-19: A Systematic Review and Quantitative Analysis

**DOI:** 10.64898/2026.07.14.26357748

**Authors:** Carolyne Assed Caires Duarte, Vanessa Schnekenberg Martins Uscocovich, Ivo Misael, Príscila Diane Assed Caires Duarte, Eduarda Baranselli Sestito, Pollyanna Nicolly da Silva

## Abstract

**Objective:** To synthesize the available evidence on the association between SARS-CoV-2-related microvascular thrombosis and acute kidney injury (AKI), with emphasis on renal outcomes, mortality, and renal replacement therapy requirements.

**Methods:** This systematic review followed the PRISMA 2020 statement and was prospectively registered in PROSPERO (CRD420251132701). PubMed/MEDLINE, Scopus, and Embase were searched for systematic reviews, including meta-analyses, and umbrella reviews investigating the association between SARS-CoV-2-related microvascular thrombosis and acute kidney injury. Two reviewers independently performed study selection, data extraction, and methodological quality assessment using AMSTAR-2 and ROBIS. Evidence was synthesized through a structured narrative synthesis supported by quantitative data extracted from the included reviews.

**Results:** Six evidence syntheses evaluating kidney involvement, thrombotic events, and microvascular mechanisms in COVID-19 were included. AKI incidence was 9.2% (95%CI 4.6–13.9) among hospitalized patients and 32.6% (95%CI 8.5–56.6) among critically ill patients. In children with multisystem inflammatory syndrome associated with SARS-CoV-2, AKI incidence was 20% (95%CI 14–28). Microvascular or thrombotic events were associated with adverse renal outcomes (OR 2.14; 95%CI 1.32– 3.48). AKI was associated with increased mortality (OR 4.68; 95%CI 1.06–20.70) and greater likelihood of renal replacement therapy requirement (OR 2.87; 95%CI 1.45– 5.68). The certainty of evidence ranged from moderate to high for the principal outcomes.

**Conclusion:** Current evidence supports an important association between microvascular thrombotic injury and COVID-19-associated AKI. These findings reinforce the relevance of endothelial dysfunction and thromboinflammatory pathways in kidney involvement during COVID-19 and highlight the need for early renal monitoring, risk stratification, and kidney-protective strategies in high-risk patients.

## Introduction

Coronavirus disease 2019 (COVID-19), caused by severe acute respiratory syndrome coronavirus 2 (SARS-CoV-2), is a multisystem disorder associated with significant morbidity and mortality. Although initially characterized as a respiratory illness, accumulating evidence has demonstrated substantial extrapulmonary involvement, particularly affecting the cardiovascular, neurological, hematological, and renal systems. Among these complications, acute kidney injury (AKI) has emerged as one of the most frequent and clinically relevant manifestations, contributing to prolonged hospitalization, increased need for intensive care, and higher mortality rates.

The mechanisms underlying COVID-19-associated AKI are complex and multifactorial. Proposed pathways include direct viral invasion of renal tissues through angiotensin-converting enzyme 2 (ACE2) receptors, systemic inflammatory responses, cytokine-mediated injury, hemodynamic instability, hypoxemia, and nephrotoxic exposures. However, increasing attention has been directed toward the role of endothelial dysfunction and microvascular thrombosis as central contributors to renal injury in patients with COVID-19.

SARS-CoV-2 infection promotes a prothrombotic state characterized by endothelial activation, complement dysregulation, platelet aggregation, and excessive inflammatory signaling. These processes contribute to thromboinflammation and the formation of microvascular thrombi in multiple organs, including the kidneys. Histopathological studies have demonstrated evidence of endothelial injury, capillary congestion, fibrin deposition, and thrombotic microangiopathy in renal tissue obtained from patients with severe COVID-19. Such findings support the hypothesis that microvascular thrombosis may impair renal perfusion, exacerbate ischemic injury, and accelerate the progression of AKI. Despite the growing number of evidence syntheses addressing COVID-19- associated AKI, no previous review has specifically focused on integrating evidence regarding the contribution of microvascular thrombosis and thromboinflammatory mechanisms to renal injury across different evidence syntheses.

Several systematic reviews and meta-analyses have investigated the incidence, prognostic implications, and pathophysiological mechanisms of AKI in COVID-19. These studies have consistently reported associations between renal dysfunction and adverse clinical outcomes, including increased mortality and greater need for renal replacement therapy. Nevertheless, the extent to which microvascular thrombosis contributes to these outcomes remains incompletely characterized, and available evidence is dispersed across multiple evidence syntheses with varying methodologies and populations.

A comprehensive synthesis of these findings is important to clarify the relationship between microvascular thrombosis and COVID-19-associated AKI, identify consistent patterns across studies, and provide clinically relevant insights for risk stratification and patient management. Understanding the role of thromboinflammatory mechanisms may also contribute to the development of preventive and therapeutic strategies aimed at reducing renal complications in patients with COVID-19.

Therefore, the present study aimed to systematically review and quantitatively synthesize evidence from systematic reviews, meta-analyses, and umbrella reviews evaluating the association between SARS-CoV-2-related microvascular thrombosis and acute kidney injury, as well as its impact on mortality, renal replacement therapy requirements, and renal function outcomes.

## Methods

This systematic review was conducted to investigate the association between SARS-CoV-2-related microvascular thrombosis and acute kidney injury (AKI), including its relationship with mortality, renal replacement therapy (RRT), and the underlying thromboinflammatory mechanisms. The review was conducted in accordance with the Preferred Reporting Items for Systematic Reviews and Meta-Analyses (PRISMA 2020) statement, and the study protocol was prospectively registered in the International Prospective Register of Systematic Reviews (PROSPERO) under registration number CRD420251132701.

The review question was developed according to the Population, Exposure, Comparison, and Outcomes (PICO) framework. The population comprised patients with confirmed COVID-19, and the exposure of interest included microvascular thrombosis, thrombotic microangiopathy, endothelial dysfunction, and other SARS-CoV-2-associated thromboinflammatory mechanisms potentially involved in kidney injury. Comparisons, when available in the included studies, involved patients with and without thrombotic involvement or different degrees of thrombotic burden. The primary outcome was the occurrence of AKI, whereas secondary outcomes included mortality, renal replacement therapy requirements, renal function outcomes, and mechanistic evidence supporting the association between microvascular thrombosis and COVID-19- associated kidney injury. As this review synthesized evidence exclusively from previously published studies, institutional ethics committee approval was not required. A comprehensive electronic literature search was conducted in PubMed/MEDLINE, Scopus, and Embase to identify systematic reviews, including meta-analyses, and umbrella reviews investigating the association between COVID-19, microvascular thrombosis, and acute kidney injury. Searches included studies published between January 2020 and March 2025, and the searches were completed in July 2025.

Search strategies combined controlled vocabulary (Medical Subject Headings [MeSH] and Emtree, when applicable) with free-text terms related to COVID-19, SARS-CoV-2, acute kidney injury, renal replacement therapy, microvascular thrombosis, thrombotic microangiopathy, and evidence synthesis study designs. Boolean operators ("AND" and "OR") were used to combine search terms, and the syntax was adapted to the indexing requirements of each database.

Records retrieved from all databases were exported for duplicate identification before eligibility assessment. The reference lists of the included studies were manually screened to identify additional eligible publications. The complete electronic search strategies, including database-specific search strings, applied filters, search dates, and the number of retrieved records, are provided in Supplementary Material S1, in accordance with the PRISMA-S recommendations.

Eligibility criteria were established a priori according to the PICO framework. Systematic reviews, systematic reviews including meta-analyses, and umbrella reviews investigating the association between COVID-19-related thromboinflammatory or microvascular mechanisms and renal outcomes were considered eligible. Evidence syntheses including patients of any age, sex, ethnicity, geographic location, or disease severity were eligible, provided that they reported at least one predefined renal outcome.

The primary outcome was acute kidney injury. Secondary outcomes included mortality, renal replacement therapy, renal function parameters, histopathological evidence of renal microvascular injury, and other clinically relevant renal outcomes associated with COVID-19. Narrative reviews, editorials, conference abstracts, expert opinions, protocols, case reports, case series, experimental animal studies, in vitro studies, and publications without relevant quantitative outcome data were excluded. Duplicate records were removed before screening, and, when overlapping evidence syntheses were identified, the most comprehensive and methodologically robust review was retained.

Following duplicate removal, two reviewers independently screened all retrieved records in two sequential stages according to the predefined eligibility criteria. First, titles and abstracts were assessed to identify potentially relevant studies. Subsequently, the full texts of all eligible publications were independently evaluated to confirm study eligibility. Disagreements regarding study selection were resolved through discussion until consensus was reached, guided by the predefined inclusion and exclusion criteria. The study selection process was documented using the PRISMA 2020 flow diagram, including the number of records identified, screened, excluded, and included at each stage.

Data extraction was independently performed by two reviewers using a standardized data extraction form. The following information was collected from each included study: first author, year of publication, country, study design, number of included primary studies, sample size, characteristics of the study population, renal outcomes, mortality, renal replacement therapy requirements, reported effect measures, methodological characteristics, and the main conclusions. Discrepancies were resolved through discussion until consensus was achieved. When overlapping evidence was identified among systematic reviews, only the most comprehensive and methodologically robust review was retained.

The methodological quality and risk of bias of the included evidence syntheses were independently assessed by two reviewers. AMSTAR-2 (A MeaSurement Tool to Assess Systematic Reviews 2) was used to evaluate systematic reviews, including those with meta-analyses, whereas ROBIS (Risk Of Bias In Systematic Reviews) was applied to umbrella reviews, when appropriate. Both instruments were selected because they are internationally recognized and specifically developed to assess the methodological quality and risk of bias of evidence synthesis studies. Disagreements between reviewers were resolved through discussion until consensus was achieved. The methodological quality assessments informed the interpretation and synthesis of the evidence.

The findings of the included reviews were synthesized using a structured narrative approach supported by quantitative data extracted from the included systematic reviews and umbrella reviews. Evidence was compared with emphasis on the incidence of acute kidney injury, mortality, renal replacement therapy requirements, renal function outcomes, and the reported mechanisms linking SARS-CoV-2-associated microvascular thrombosis to kidney injury. Reported effect estimates and corresponding 95% confidence intervals for the principal renal outcomes were extracted directly from the included evidence syntheses and graphically summarized to facilitate comparison across outcomes. No de novo meta-analysis, statistical pooling, or recalculation of pooled effect estimates was performed. The direction, consistency, and strength of the reported findings were interpreted descriptively while considering the methodological quality of each review. When overlapping evidence was identified, priority was given to the most comprehensive and methodologically robust review to minimize duplication of primary studies and avoid overrepresentation of the same evidence. Methodological quality assessments obtained using AMSTAR-2 and ROBIS informed data interpretation, giving greater weight to reviews with lower risk of bias and higher methodological quality. Sources of heterogeneity reported by the included reviews were also considered during interpretation of the findings.

When overlapping evidence was identified, priority was given to the most comprehensive and methodologically robust review to minimize duplication of primary studies and avoid overrepresentation of the same evidence. Methodological quality assessments obtained using AMSTAR-2 and ROBIS were considered during data interpretation, giving greater weight to reviews with lower risk of bias and higher methodological quality. Quantitative effect estimates reported by the included systematic reviews were extracted and descriptively summarized without performing a new meta-analysis or recalculation of pooled effect sizes. Sources of heterogeneity reported by the included reviews were considered during evidence interpretation.

The certainty of evidence for the principal outcomes was evaluated using the GRADE (Grading of Recommendations Assessment, Development and Evaluation) approach. The assessment considered the domains of risk of bias, inconsistency, indirectness, imprecision, publication bias, and magnitude of effect, and the overall certainty ratings were used to support interpretation of the findings (Supplementary Table S1).

## Results

The electronic search conducted in PubMed/MEDLINE, Scopus, and Embase identified 613 records. After removal of one duplicate, 612 records underwent title and abstract screening. Of these, 455 records were excluded because they did not meet the predefined eligibility criteria. A total of 157 reports were assessed for eligibility, of which 111 were excluded because they were unrelated to renal outcomes, had an editorial design, or did not meet the methodological scope of the review. Among the remaining 46 reports, 40 were further excluded because they were not systematic reviews or meta-analyses (n = 15), had abstracts inconsistent with the objective of the study (n = 20), or presented full texts incompatible with the inclusion criteria (n = 5). Ultimately, six studies met all eligibility criteria and were included in the qualitative and quantitative synthesis. The study selection process is shown in the PRISMA 2020 flow diagram (Figure 1).

**Figure 1.**
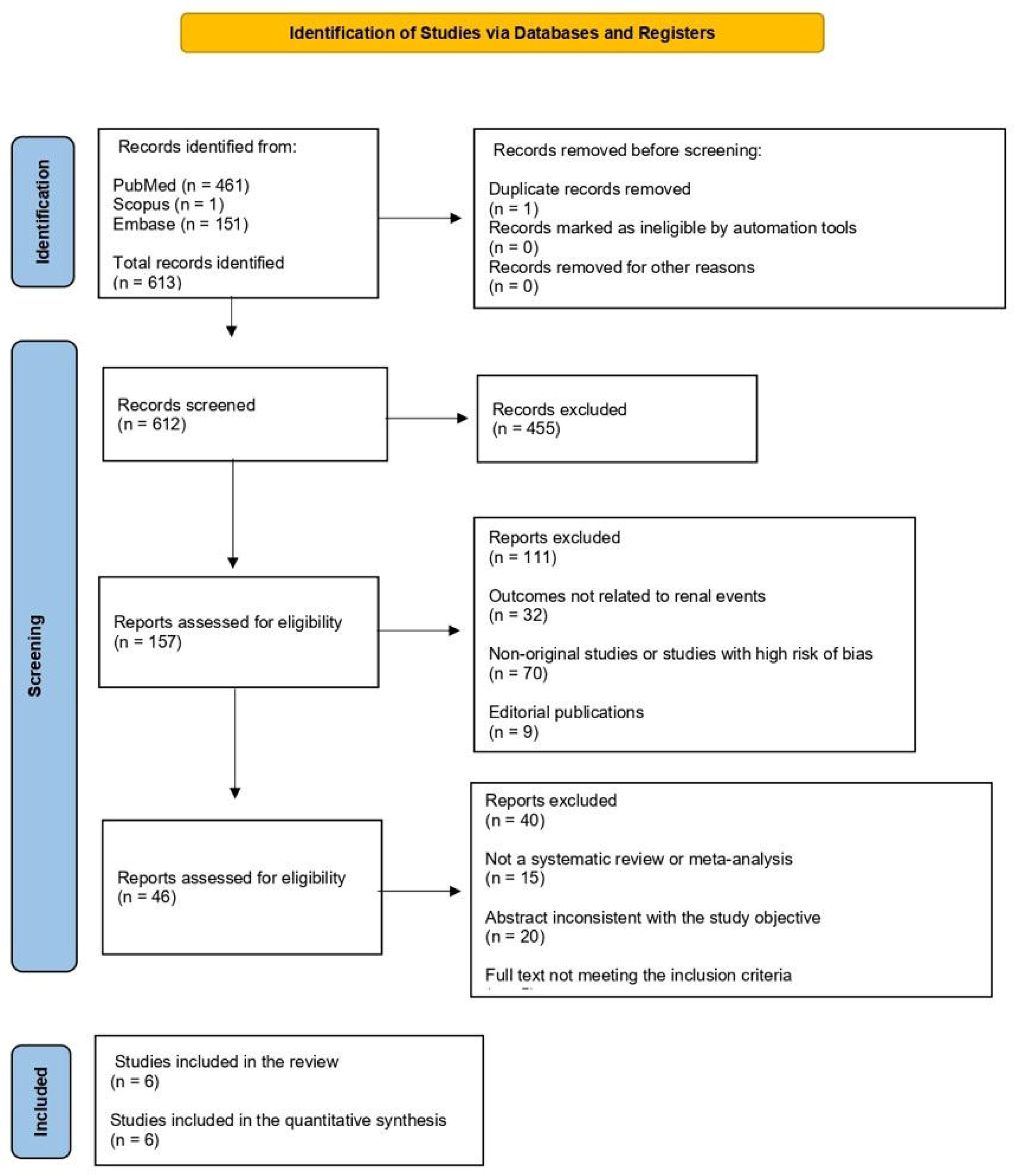
PRISMA 2020 flow diagram illustrating the identification, screening, eligibility assessment, and inclusion of studies in the systematic review and quantitative synthesis.

The six included studies were published between 2020 and 2023 and comprised systematic reviews, systematic reviews with meta-analysis, and umbrella reviews evaluating renal outcomes, thrombotic events, and microvascular mechanisms associated with COVID-19. The included evidence syntheses addressed adult hospitalized patients, critically ill patients admitted to intensive care units, and pediatric patients with multisystem inflammatory syndrome in children (MIS-C). The main characteristics and findings of the included studies are summarized in Table 1.

**Table 1.**
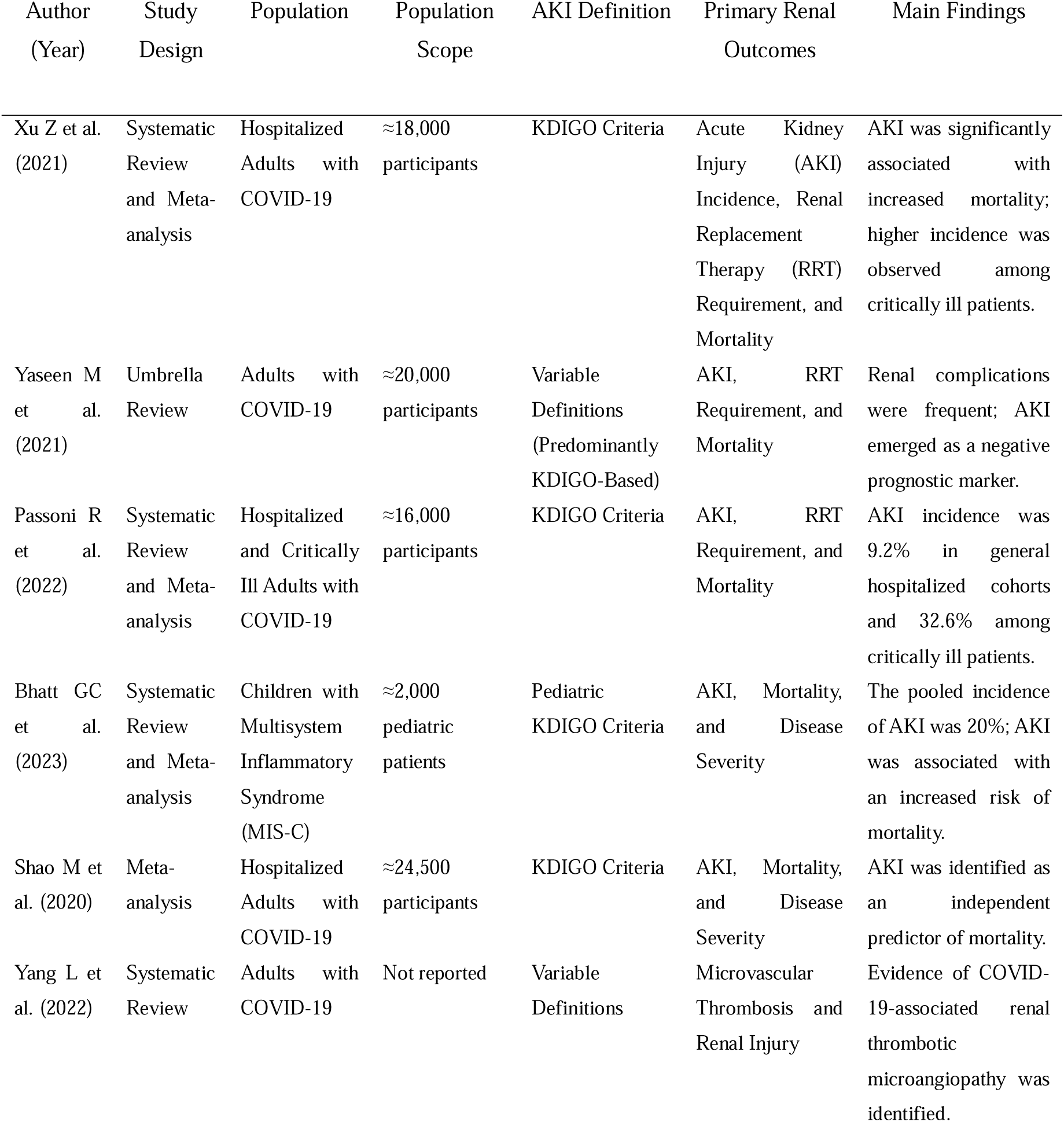
Characteristics of the systematic reviews included in the quantitative synthesis.

| Author<br>(Year) | Study<br>Design | Population | Population<br>Scope | AKI Definition | Primary Renal<br>Outcomes | Main Findings |
| --- | --- | --- | --- | --- | --- | --- |
| Xu Z et al.<br>(2021) | Systematic<br>Review<br>and Meta-analysis | Hospitalized<br>Adults with<br>COVID-19 | ≈18,000<br>participants | KDIGO Criteria | Acute Kidney<br>Injury (AKI)<br>Incidence, Renal<br>Replacement<br>Therapy (RRT)<br>Requirement, and<br>Mortality | AKI was significantly<br>associated with<br>increased mortality;<br>higher incidence was<br>observed among<br>critically ill patients. |
| Yaseen M<br>et al.<br>(2021) | Umbrella<br>Review | Adults with<br>COVID-19 | ≈20,000<br>participants | Variable<br>Definitions<br>(Predominantly<br>KDIGO-Based) | AKI, RRT<br>Requirement, and<br>Mortality | Renal complications<br>were frequent; AKI<br>emerged as a negative<br>prognostic marker. |
| Passoni R<br>et al.<br>(2022) | Systematic<br>Review<br>and Meta-analysis | Hospitalized<br>and Critically<br>Ill Adults with<br>COVID-19 | ≈16,000<br>participants | KDIGO Criteria | AKI, RRT<br>Requirement, and<br>Mortality | AKI incidence was<br>9.2% in general<br>hospitalized cohorts<br>and 32.6% among<br>critically ill patients. |
| Bhatt GC<br>et al.<br>(2023) | Systematic<br>Review<br>and Meta-analysis | Children with<br>Multisystem<br>Inflammatory<br>Syndrome<br>(MIS-C) | ≈2,000<br>pediatric<br>patients | Pediatric<br>KDIGO Criteria | AKI, Mortality,<br>and Disease<br>Severity | The pooled incidence<br>of AKI was 20%; AKI<br>was associated with<br>an increased risk of<br>mortality. |
| Shao M et<br>al. (2020) | Meta-analysis | Hospitalized<br>Adults with<br>COVID-19 | ≈24,500<br>participants | KDIGO Criteria | AKI, Mortality,<br>and Disease<br>Severity | AKI was identified as<br>an independent<br>predictor of mortality. |
| Yang L et<br>al. (2022) | Systematic<br>Review | Adults with<br>COVID-19 | Not reported | Variable<br>Definitions | Microvascular<br>Thrombosis and<br>Renal Injury | Evidence of COVID-19-associated renal<br>thrombotic<br>microangiopathy was<br>identified. |

Acute kidney injury (AKI) definitions were predominantly based on Kidney Disease: Improving Global Outcomes (KDIGO) criteria, although some variability was observed across studies. The most frequently reported comorbidities among patients with AKI included hypertension, diabetes mellitus, and pre-existing chronic kidney disease.

The incidence of AKI varied according to clinical setting and disease severity. In general hospitalized cohorts, pooled AKI incidence was 9.2% (95% CI 4.6–13.9). Among critically ill patients, AKI incidence was substantially higher, reaching 32.6% (95% CI 8.5–56.6). In pediatric populations with MIS-C, the pooled incidence of AKI was 20% (95% CI 14–28). The requirement for renal replacement therapy (RRT) ranged from 3.2% to 15% across the included evidence syntheses. Mortality among patients with AKI showed wide variability, ranging from 17.0% to 83.9%, with a pooled estimate of 50.4% (95% CI 17.0–83.9).

Quantitative synthesis demonstrated that AKI was significantly associated with adverse clinical outcomes in patients with COVID-19. The presence of AKI was associated with increased mortality (OR 4.68; 95%CI 1.06–20.70) and a greater likelihood of requiring renal replacement therapy (OR 2.87; 95%CI 1.45–5.68). Microvascular or thrombotic events were significantly associated with unfavorable renal outcomes, with a pooled odds ratio of 2.14 (95%CI 1.32–3.48). These reported effect estimates were derived from aggregate effect measures reported across the included evidence syntheses. For the principal renal outcomes, the effect estimates reported by the included systematic reviews were graphically summarized to facilitate comparison across outcomes (Figure 2). The reported odds ratios and corresponding 95% confidence intervals were extracted directly from the included evidence syntheses, without recalculation or statistical pooling. Because the reported effect estimates originated from previously published evidence syntheses with different populations, inclusion criteria, and analytical approaches, the findings were interpreted descriptively, considering the consistency and direction of the reported associations rather than generating new pooled estimates. Heterogeneity reported by the included reviews was considered during interpretation because the included reviews differed in population characteristics, AKI definitions, disease severity, and outcome reporting.

**Figure 2.**
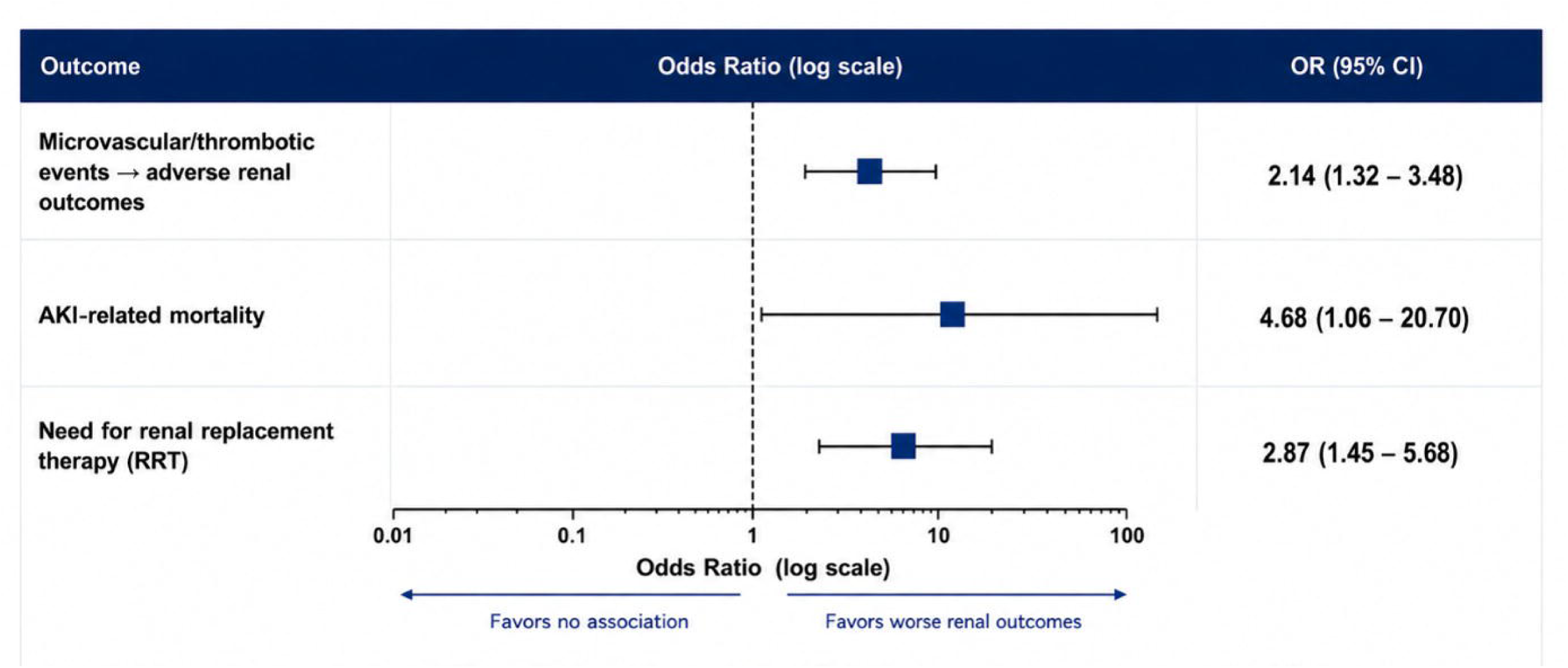
Summary plot of reported effect estimates for the principal renal outcomes associated with COVID-19.

The methodological quality of the included evidence syntheses was assessed using AMSTAR-2, and the certainty of evidence for the principal outcomes was evaluated using the GRADE framework (Table S1). Overall, the certainty of evidence ranged from moderate to high across the principal outcomes, with greater confidence observed for the major dichotomous outcomes, including AKI, mortality, and RRT requirement.

## Discussion

The findings of this systematic review and quantitative synthesis reinforce that acute kidney injury (AKI) is one of the most clinically relevant extrapulmonary complications of COVID-19 and should be interpreted within the broader framework of systemic endothelial dysfunction and thromboinflammation. Across the included evidence syntheses, AKI was consistently associated with increased mortality, greater renal replacement therapy (RRT) requirements, and worse clinical outcomes. Importantly, thrombotic and microvascular events were also associated with unfavorable renal outcomes, supporting the biological plausibility that renal microcirculatory injury contributes substantially to COVID-19-associated kidney dysfunction.

Since the early stages of the pandemic, it has become increasingly clear that COVID-19 extends beyond respiratory involvement and affects multiple organ systems through complex inflammatory and vascular pathways. SARS-CoV-2 infection has been associated with endothelial activation, complement dysregulation, platelet activation, and a sustained prothrombotic state. Within the kidney, these alterations may compromise microcirculatory integrity through glomerular and peritubular microvascular injury, impairing tissue perfusion and favoring tubular dysfunction. Histopathological reports describing endothelial injury, fibrin deposition, capillary congestion, renal infarction, and thrombotic microangiopathy provide biological plausibility for the associations observed in the present review.

The substantial variation in AKI incidence according to disease severity further highlights the multifactorial nature of renal involvement in COVID-19. While pooled incidence remained relatively lower among general hospitalized populations, rates increased considerably among critically ill patients. This pattern likely reflects the combined effects of systemic inflammation, hemodynamic instability, hypoxemia, endothelial dysfunction, coagulation abnormalities, and microvascular injury. Rather than acting independently, these mechanisms appear to amplify one another, creating a biological environment particularly favorable to renal injury and disease progression.

From a nephrology practice perspective, these findings reinforce the importance of early and repeated renal assessment in patients with moderate-to-severe COVID-19, particularly among those admitted to intensive care units or presenting with markers of systemic inflammation, coagulation activation, hemodynamic instability, or pre-existing kidney disease. Standardized AKI definitions, especially KDIGO criteria, remain essential for early diagnosis, staging, prognostic assessment, and comparability across clinical studies. Routine monitoring of serum creatinine, urine output, and other KDIGO-recommended parameters may facilitate earlier recognition of AKI and timely nephrology consultation. The consistent association between AKI and mortality observed across evidence syntheses supports the need for structured renal monitoring protocols in high-risk patients.

Particularly noteworthy was the strong association between AKI and mortality. The magnitude of the observed effect suggests that AKI should not be viewed solely as a marker of disease severity, but also as a clinically meaningful component of the pathophysiological cascade associated with multiorgan dysfunction. Acute impairment of kidney function may contribute to fluid overload, electrolyte disturbances, acid-base imbalance, accumulation of inflammatory mediators, and worsening systemic homeostasis, all of which may further compromise organ function in critically ill patients. This interpretation is consistent with previous evidence syntheses in which AKI emerged as one of the strongest predictors of mortality among hospitalized patients with COVID-19.

The association between AKI and increased RRT requirements has direct implications for nephrology services and critical care planning. During the most critical phases of the pandemic, many healthcare systems experienced increased demand for dialysis resources, particularly in intensive care settings. The present findings support the need for early nephrology involvement when renal function deteriorates, careful fluid and hemodynamic management, avoidance of nephrotoxic exposures whenever possible, and timely preparation for kidney replacement therapy in patients at higher risk of progression.

An important aspect of the present review is the inclusion of pediatric evidence, particularly studies involving multisystem inflammatory syndrome in children (MIS-C). Although children generally experience lower mortality rates than adults, AKI was not uncommon among patients with MIS-C and was associated with clinically significant adverse outcomes. The intense inflammatory response characteristic of MIS-C may promote endothelial activation and microvascular injury through mechanisms that partially overlap with those observed in severe adult disease. These findings highlight the importance of maintaining vigilance for renal complications across different age groups and clinical presentations.

The association observed between thrombotic and microvascular events and unfavorable renal outcomes deserves particular emphasis. The pooled estimates demonstrated that microvascular involvement was associated with more than a twofold increase in the likelihood of adverse renal outcomes. In this context, renal thrombotic microangiopathy may represent an important pathophysiological link connecting endothelial injury, coagulation abnormalities, and kidney dysfunction. The interaction between viral-mediated endothelial damage and activation of thromboinflammatory pathways may help explain why some patients develop severe renal complications even in the absence of overt macrovascular thrombosis or profound systemic hypotension.

From a mechanistic perspective, the interaction between SARS-CoV-2 and angiotensin-converting enzyme 2 (ACE2) receptors may contribute directly and indirectly to renal injury through endothelial dysfunction, apoptosis, inflammatory activation, and exposure of procoagulant factors. The resulting heterogeneity of renal perfusion may explain the discrepancy occasionally observed between extensive structural injury and preserved global renal function during the early stages of disease. Taken together, these observations support the characterization of severe COVID-19 as a systemic thromboinflammatory disorder with important microvascular renal manifestations.

The findings reported here are broadly consistent with previous systematic reviews and meta-analyses describing elevated rates of AKI, increased mortality, and greater use of RRT among patients with COVID-19. However, the present study expands current knowledge by specifically emphasizing the contribution of microvascular injury and thrombotic mechanisms to renal dysfunction and by integrating evidence from systematic reviews, meta-analyses, and umbrella reviews. This approach provides a broader perspective on the interaction between endothelial dysfunction, thrombosis, and kidney injury throughout the clinical spectrum of COVID-19.

The heterogeneity observed across studies likely reflects differences in patient populations, disease severity, healthcare settings, diagnostic criteria for AKI, and temporal phases of the pandemic. Such variability is expected in evidence derived from multiple countries and healthcare systems and should be considered when interpreting pooled estimates. Nevertheless, the consistency of the principal associations across diverse populations supports the overall robustness of the findings.

Several methodological strengths enhance the reliability of the present review. The study protocol was prospectively registered in PROSPERO, the review was conducted according to PRISMA 2020 recommendations, and a comprehensive search strategy was applied across multiple databases. Methodological quality was assessed using AMSTAR-2, while certainty of evidence was evaluated through the GRADE framework. Furthermore, the consistency of findings across the included evidence syntheses, together with their methodological quality, supports the robustness of the principal conclusions.

The translational relevance of these findings lies in the recognition that COVID-19- associated AKI may reflect not only systemic illness severity but also a potentially targetable microvascular phenotype. Endothelial injury, complement activation, platelet activation, and coagulation abnormalities may interact to impair renal perfusion and promote tubular injury. Better characterization of these pathways may help identify biomarkers of renal risk, guide individualized anticoagulation or anti-inflammatory strategies, and support future trials focused on prevention of microvascular kidney injury.

Some limitations should be acknowledged. First, the analysis was based on previously published evidence syntheses rather than individual patient-level data. Consequently, potential overlap of primary studies across the included systematic reviews represents an inherent limitation of overview-level evidence synthesis and was considered during interpretation of pooled findings rather than treated as independent observations. Second, differences in study design, patient characteristics, disease severity, and outcome definitions may have contributed to residual heterogeneity. Third, much of the available evidence originated during earlier phases of the pandemic and may not fully reflect the influence of emerging viral variants, vaccination, and evolving therapeutic strategies. Finally, although the available evidence consistently supports an association between microvascular injury and AKI, the observational nature of the underlying data precludes definitive causal inference.

Taken together, the available evidence suggests that AKI in COVID-19 represents more than an isolated renal complication and should be considered a clinically relevant manifestation of systemic thromboinflammatory injury in which endothelial dysfunction and microvascular damage play central roles. Improved understanding of these mechanisms may contribute to earlier identification of high-risk patients, more effective risk stratification, and the development of targeted therapeutic strategies aimed at mitigating renal and systemic complications associated with COVID-19.

## Conclusion

This systematic review and quantitative synthesis demonstrates that acute kidney injury is a frequent and clinically relevant complication of COVID-19 and is consistently associated with increased mortality, greater need for renal replacement therapy, and worse clinical outcomes. The available evidence further supports a significant association between microvascular thrombotic injury and adverse renal outcomes, reinforcing the central role of endothelial dysfunction and thromboinflammatory pathways in the pathogenesis of COVID-19-associated kidney injury.

The available evidence suggests that renal involvement in COVID-19 should be understood within the broader context of systemic microvascular and thromboinflammatory injury rather than as an isolated organ complication. Recognition of these mechanisms has important clinical implications, emphasizing the value of early identification of renal dysfunction, careful monitoring of kidney function and coagulation parameters, and timely implementation of kidney-protective strategies in high-risk patients.

Collectively, these findings suggest that microvascular thrombosis constitutes an important biological pathway linking SARS-CoV-2 infection to kidney injury and adverse clinical outcomes. Further studies are warranted to clarify causal mechanisms and to evaluate targeted interventions aimed at reducing endothelial and microvascular damage in patients with COVID-19.

## Data Availability

No new data were generated in this study. All data analyzed were extracted from previously published studies identified through systematic searches of publicly available bibliographic databases. All data supporting the findings of this study are included within the manuscript and its supplementary materials.

## Acknowledgements

The authors have no acknowledgements to declare.

## Ethical Approval

Ethics committee approval was not required because this study is a systematic review and quantitative synthesis of previously published studies and did not involve direct participation of human subjects or access to identifiable individual patient data.

## Consent to Participate

Not applicable. This study analyzed data from previously published studies and did not involve direct recruitment of participants.

## Author Contributions (CRediT)

Carolyne Assed Caires Duarte: Conceptualization, Methodology, Investigation, Data Curation, Formal Analysis, Visualization, Project Administration, Writing – Original Draft, Writing – Review & Editing.

Vanessa Schnekenberg Martins Uscocovich: Methodology, Validation, Supervision, Resources, Writing – Review & Editing.

Ivo Misael: Methodology, Validation, Supervision, Resources, Writing – Review & Editing.

Príscila Diane Assed Caires Duarte: Methodology, Validation, Supervision, Resources, Writing – Review & Editing.

Eduarda Baranselli Sestito: Investigation, Data Curation, Visualization.

Pollyanna Nicolly da Silva: Investigation, Data Curation, Visualization.

All authors contributed to the interpretation of data, critically reviewed the manuscript, approved the final version, and agree to be accountable for all aspects of the work.

## Data Availability Statement

No new data were generated in this study. All data analyzed were extracted from previously published studies and are included within the article and its supplementary materials.

## AI Statement

Artificial intelligence tools were used exclusively to improve language and editorial clarity. All scientific content, interpretation, critical revision, and final approval were performed by the authors, who assume full responsibility for the manuscript.

---

**Table S1.** Assessment of Certainty of Evidence According to the GRADE Framework for the Principal Renal Outcomes Associated with COVID-19.

| Domain | Assessment | Summary Justification | Certainty Rating |
| --- | --- | --- | --- |
| Risk of Bias | Low | Studies were classified as having moderate-to-high methodological quality according to the adopted assessment tools and demonstrated overall consistency across quality domains. | No serious concerns |
| Inconsistency | Not Serious | Clinical and methodological heterogeneity was expected across the included evidence syntheses because of differences in populations, AKI definitions, disease severity, and outcome reporting; however, the direction of effects was generally consistent across the principal outcomes. | No serious concerns |
| Imprecision | Not Serious | Relatively narrow 95% confidence intervals were observed for the principal outcomes, including acute kidney injury and mortality. | No serious concerns |
| Publication Bias | Low | Low likelihood of publication bias, supported by a non-significant Egger's regression test ($p = 0.123$) and visual symmetry of the funnel plots. | No serious concerns |
| Indirectness | Not Serious | Population, exposure, and outcomes were directly related to microvascular thrombosis and COVID-19-associated kidney injury. | No serious concerns |
| Magnitude of Effect | Large Effect | Clinically relevant associations were observed, with odds ratios exceeding 3.0 for acute kidney injury and mortality outcomes. | Upgraded |
| Overall Certainty of Evidence | Moderate to High | Overall certainty of evidence was rated as moderate-to-high across the principal outcomes. | Moderate to High |

