## Supplementary Material S1 for "Microvascular Thrombosis and Acute Kidney Injury in COVID-19: A Systematic Review and Quantitative Analysis": SUPLEMENTARY MATERIAL S1.docx

**Electronic Search Strategies**

A comprehensive literature search was performed in **PubMed/MEDLINE, Scopus, and Embase** to identify systematic reviews, including meta-analyses, and umbrella reviews investigating the association between SARS-CoV-2 infection, microvascular thrombosis, and acute kidney injury. Searches covered studies published between **January 2020 and March 2025**. All electronic searches were completed in **July 2025**. Controlled vocabulary (**MeSH** and **Emtree**, when applicable) and free-text terms were combined using Boolean operators ("AND" and "OR"), with syntax adapted to the indexing requirements of each database. Database-specific syntax was adapted without changing the conceptual search framework.

**Electronic Search Strategies**

PubMed/MEDLINE

**Search strategy 1**

microvascular thrombosis AND kidney

**Search strategy 2**

("umbrella review") AND AKI AND COVID-19

**Search strategy 3**

("primary studies") AND ("systematic review" OR "meta-analysis") AND AKI AND COVID-19

Filter: Free Full Text

Records retrieved: 461

Scopus

Search strategy

TITLE-ABS-KEY(

("Acute Kidney Injury" OR AKI)

AND

("Continuous Renal Replacement Therapy" OR CRRT OR "Renal Replacement Therapy")

AND

(COVID-19 OR SARS-CoV-2)

AND

("Systematic Review" OR "Meta-analysis")

)

Records retrieved: 1

Duplicate removed before screening:1

Embase

Search strategy

("Acute Kidney Injury" OR AKI)

AND

("Continuous Renal Replacement Therapy" OR CRRT OR "Renal Replacement Therapy")

AND

(COVID-19 OR SARS-CoV-2)

AND

("Systematic Review" OR "Meta-analysis")

Records retrieved: 151

Total records identified: 613

Records screened after duplicate removal: 612
